## Supplemental Tables 1 to 20 for "Active vaccine safety surveillance: Experience from a prospective cohort event monitoring study of COVID-19 vaccines in Kenya"

**Supplementary Table 1.** Sample sizes needed to rule out various levels of an increase in the risk of adverse events of special interest (AESI) if no event is observed within a 42-day risk window. Cells with a sample size of ≤10,000 are shaded. The calculation of the 95% CI if no event is observed is based on the methods proposed by Eypasch et al., 1995.

| **AESI** | **Background rate per 100,000 person years** | **Relative risk** | **Theoretical rate per 100,000 person years assuming increased risk (theoretical 95% CI)** | **Sample size** | **95% CI per 100,000 person years if no event is observed** |
| --- | --- | --- | --- | --- | --- |
| ARDS | 39 | 1.5 | **59** (45, 76) | 59,370 | 0, 44 |
|  |  | 2 | **78** (62, 97) | 42,983 | 0, 61 |
|  |  | 3 | **117** (97, 140) | 27,225 | 0, 96 |
|  |  | 4 | **156** (132, 182) | 19,829 | 0, 131 |
|  |  | 5 | **195** (169, 224) | 15,557 | 0, 168 |
|  | 90 | 1.5 | **135** (113, 160) | 23,239 | 0, 112 |
|  |  | 2 | **180** (155, 208) | 16,967 | 0, 154 |
|  |  | 3 | **270** (239, 304) | 10,966 | 0, 238 |
|  |  | 4 | **360** (324, 399) | 8,077 | 0, 323 |
|  |  | 5 | **450** (409, 494) | 6,384 | 0, 408 |
|  | 150 | 1.5 | **225** (197, 256) | 13,332 | 0, 196 |
|  |  | 2 | **300** (267, 336) | 9,801 | 0, 266 |
|  |  | 3 | **450** (409, 494) | 6,384 | 0, 408 |
|  |  | 4 | **600** (553, 650) | 4,724 | 0, 552 |
|  |  | 5 | **750** (697, 806) | 3,744 | 0, 696 |
|  | 193 | 1.5 | **290** (258, 325) | 10,161 | 0, 257 |
|  |  | 2 | **386** (348, 426) | 7,504 | 0, 347 |
|  |  | 3 | **579** (533, 628) | 4,903 | 0, 532 |
|  |  | 4 | **772** (718, 828) | 3,634 | 0, 717 |
|  |  | 5 | **965** (905, 1028) | 2,884 | 0, 904 |
| Thrombosis | 80 | 1.5 | **120** (99, 143) | 26,471 | 0, 98 |
|  |  | 2 | **160** (136, 187) | 19,288 | 0, 135 |
|  |  | 3 | **240** (211, 272) | 12,439 | 0, 210 |
|  |  | 4 | **320** (286, 357) | 9,151 | 0, 285 |
|  |  | 5 | **400** (362, 441) | 7,227 | 0, 361 |
| Acute aseptic arthritis | 100 | 1.5 | 150 (127, 176) | 20,699 | 0, 126 |
|  |  | 2 | 200 (173, 230) | 15,137 | 0, 172 |
|  |  | 3 | 300 (267, 336) | 9,801 | 0, 266 |
|  |  | 4 | 400 (362, 441) | 7,227 | 0, 361 |
|  |  | 5 | 500 (457, 546) | 5,716 | 0, 456 |
|  | 500 | 1.5 | 750 (697, 806) | 3,744 | 0, 696 |
|  |  | 2 | 1000 (939, 1064) | 2,780 | 0, 938 |
|  |  | 3 | 1500 (1425, 1578) | 1,831 | 0, 1,424 |
|  |  | 4 | 2000 (1913, 2090) | 1,363 | 0, 1,912 |
|  |  | 5 | 2500 (2403, 2600) | 1,085 | 0, 2,402 |
|  | 1000 | 1.5 | 1500 (1425, 1578) | 1,831 | 0, 1,424 |
|  |  | 2 | 2000 (1913, 2090) | 1,363 | 0, 1,912 |
|  |  | 3 | 3000 (2894, 3109) | 901 | 0, 2,893 |
|  |  | 4 | 4000 (3877, 4126) | 673 | 0, 3,876 |
|  |  | 5 | 1500 (1425, 1578) | 536 | 0, 4,861 |
|  | 1600 | 1.5 | 2400 (2305, 2498) | 1,132 | 2,304 |
|  |  | 2 | 3200 (3090, 3313) | 844 | 3,089 |
|  |  | 3 | 4800 (4665, 4938) | 559 | 4,664 |
|  |  | 4 | 6400 (6244, 6559) | 418 | 6,243 |
|  |  | 5 | 8000 (7826, 8177) | 333 | 7,825 |

**Supplementary Table 2.** Margins of error for different levels of reactogenicity prevalence for a sample size of 1,000. Margins of error are based on the width of the exact binomial 95% confidence intervals for a proportion.

| **Prevalence estimate** | **Margin of error** |
| --- | --- |
| 1% | ±0.6% |
| 2% | ±0.8% |
| 5% | ±1.3% |
| 10% | ±1.9% |
| 25% | ±2.7% |
| 50% | ±3.1% |

**Supplementary Table 3.** Distribution of vaccines received by brand and dose among enrolled participants at baseline (this excludes one re-enrolled participant with follow up over two doses).

| **Brand** | **Dose** | **Reactogenicity subset**  **N=1000**  **n (%)** | **Non-reactogenicity subset**  **N=1439**  **n (%)** | **Overall N=2439**  **n (%)** |
| --- | --- | --- | --- | --- |
| Pfizer | 1^st^ | 208 (20.8) | 267 (18.5) | 475 (19.5) |
|  | 2^nd^ | 108 (10.8) | 155 (10.8) | 263 (10.8) |
|  | 3^rd^ | 64 (6.4) | 75 (5.2) | 139 (5.7) |
|  | 4^th^ | 1 (0.1) | 2 (0.1) | 3 (0.1) |
| Johnson & Johnson | 1^st^ | 366 (36.6) | 445 (30.9) | 811 (33.3) |
|  | 2^nd^ | 87 (8.7) | 143 (9.9) | 230 (9.4) |
|  | 3^rd^ | 56 (5.6) | 64 (4.4) | 120 (4.9) |
|  | 4^th^ | 4 (0.4) | 2 (0.1) | 6 (0.2) |
| Moderna | 1^st^ | 28 (2.8) | 117 (8.1) | 145 (5.9) |
|  | 2^nd^ | 45 (4.5) | 98 (6.8) | 143 (5.9) |
|  | 3^rd^ | 28 (2.8) | 66 (4.6) | 94 (3.9) |
|  | 4^th^ | 5 (0.5) | 5 (0.3) | 10 (0.4) |

**Supplementary Table 4**. Summary of follow up periods within the cohort.

| **Study week number** | **Number of participants for whom this was the terminal observation period** | **Percent** |
| --- | --- | --- |
| 1 | 15 | 0.6 |
| 2 | 12 | 0.5 |
| 3 | 10 | 0.4 |
| 4 | 15 | 0.6 |
| 5 | 8 | 0.3 |
| 6 | 11 | 0.5 |
| 7 | 7 | 0.3 |
| 8 | 13 | 0.5 |
| 9 | 25 | 1.1 |
| 10 | 20 | 0.8 |
| 11 | 23 | 1.0 |
| 12 | 12 | 0.5 |
| 13 | 2,183 | 92.7 |
| **Total** | 2,354 | 100 |

**Supplementary Table 5.** Frequency of systemic reactogenicity at baseline (within three days prior to vaccination) and daily during the first week post-vaccination. P-values compare the frequency of the event post-vaccination to baseline. Day 0 is the day of vaccination.

|  | **Chills** | | **Fatigue** | | **Fever** | | **Headache** | |
| --- | --- | --- | --- | --- | --- | --- | --- | --- |
|  | **% (95% CI)** | **p-value** | **% (95% CI)** | **p-value** | **% (95% CI)** | **p-value** | **% (95% CI)** | **p-value** |
| **Day -3 to -1 (N= 956)** | 8.6 (6.8-10.3) | Ref | 9.4 (7.6-11.3) | Ref | 5.7 (4.3-7.2) | Ref | 22.8 (20.1-25.5) | Ref |
| **Day 0 (N= 893)** | 15.3 (13.0-17.7) | **<0.001** | 24.1 (21.3-26.9) | **<0.001** | 13.9 (11.6-16.2) | **<0.001** | 21.3 (18.6-24.0) | 0.425 |
| **Day 1 (N= 884)** | 13.1 (10.9-15.3) | **<0.001** | 25.5 (22.6-28.3) | **<0.001** | 12.0 (9.8-14.1) | **<0.001** | 19.5 (16.8-22.1) | 0.103 |
| **Day 2 (N= 885)** | 6.9 (5.2-8.6) | 0.228 | 14.5 (12.1-16.8) | **0.001** | 5.4 (3.9-6.9) | 0.834 | 12.0 (9.8-14.1) | **<0.001** |
| **Day 3 (N= 881)** | 4.8 (3.4-6.2) | **0.005** | 10.6 (8.5-12.6) | 0.292 | 2.7 (1.6-3.8) | **0.001** | 9.1 (7.2-11.0) | **<0.001** |
| **Day 4 (N= 876)** | 3.1 (1.9-4.2) | **<0.001** | 7.1 (5.4-8.8) | 0.090 | 3.7 (2.4-4.9) | **0.034** | 7.6 (5.9-9.4) | **<0.001** |
| **Day 5 (N= 874)** | 3.1 (1.9-4.2) | **<0.001** | 6.2 (4.6-7.8) | **0.018** | 2.3 (1.3-3.3) | **<0.001** | 5.3 (3.8-6.7) | **<0.001** |
| **Day 6 (N= 890)** | 2.6 (1.5-3.6) | **<0.001** | 5.4 (3.9-6.9) | **0.002** | 1.9 (1.0-2.8) | **<0.001** | 6.5 (4.9-8.1) | **<0.001** |

|  | **Joint pain** | | **Malaise** | | **Muscle aches** | | **Nausea** | |
| --- | --- | --- | --- | --- | --- | --- | --- | --- |
|  | **% (95% CI)** | **p-value** | **% (95% CI)** | **p-value** | **% (95% CI)** | **p-value** | **% (95% CI)** | **p-value** |
| **Day -3 to -1 (N= 956)** | 11.3 (9.3-13.3) | Ref | 10.2 (8.3-12.2) | Ref | 5.9 (4.4-7.3) | Ref | 4.4 (3.1-5.7) | Ref |
| **Day 0 (N= 893)** | 16.7 (14.2-19.1) | **0.001** | 20.5 (17.8-23.1) | **<0.001** | 11.5 (9.4-13.6) | **<0.001** | 7.6 (5.9-9.4) | **0.006** |
| **Day 1 (N= 884)** | 16.1 (13.6-18.5) | **0.001** | 19.7 (17.1-22.3) | **<0.001** | 11.5 (9.4-13.6) | **<0.001** | 7.9 (6.1-9.7) | **0.001** |
| **Day 2 (N= 885)** | 9.4 (7.5-11.3) | 0.129 | 9.8 (7.9-11.8) | 0.932 | 6.4 (4.8-8.1) | 0.764 | 5.8 (4.2-7.3) | 0.254 |
| **Day 3 (N= 881)** | 5.8 (4.2-7.3) | **<0.001** | 7.0 (5.3-8.7) | **0.027** | 4.4 (3.1-5.8) | 0.314 | 3.4 (2.2-4.6) | 0.313 |
| **Day 4 (N= 876)** | 4.5 (3.1-5.8) | **<0.001** | 4.5 (3.1-5.8) | **<0.001** | 3.8 (2.5-5.0) | 0.053 | 2.2 (1.2-3.1) | **0.013** |
| **Day 5 (N= 874)** | 3.2 (2.0-4.4) | **<0.001** | 3.3 (2.1-4.5) | **<0.001** | 2.2 (1.2-3.1) | **<0.001** | 2.7 (1.7-3.8) | 0.058 |
| **Day 6 (N= 890)** | 2.9 (1.8-4.0) | **<0.001** | 3.8 (2.6-5.1) | **<0.001** | 2.2 (1.3-3.2) | **<0.001** | 1.8 (0.9-2.7) | **0.001** |

**Supplementary Table 6.** Summary of systemic reactogenicity events reported within the first week of COVID-19 vaccination**.**

| **Systemic reactogenicity event** | **N=956**  **n with event (%)** | **Median duration of events in days (IQR)** |
| --- | --- | --- |
| Fatigue | 422(44.1) | 2(1-3) |
| Headache | 370(38.7) | 2(1-2) |
| Malaise | 346(36.2) | 1(1-2) |
| Joint Pain | 308(32.2) | 1(1-2) |
| Muscle Aches | 247(25.8) | 1(1-2) |
| Chills | 246(25.7) | 1(1-2) |
| Fever | 233(24.4) | 1(1-2) |
| Nausea | 171(17.9) | 1(1-2) |
| Any systemic reactogenicity event ^a^ | 595(62.2) | 2(1-3) |
| None ^b^ | 361(37.8) | .. |

Abbreviation: IQR, Interquartile range. ^a^ Any systemic reactogenicity event denotes participants who reported at least one systemic reactogenicity event. ^b^ None denotes participants who did not report any systemic reactogenicity event.

**Supplementary Table 7.** Stratification of systemic reactogenicity events by severity.

| **Systemic reactogenicity event** | **None^a^**  **n (%)** | **Mild^b^**  **n (%)** | **Moderate^c^**  **n (%)** | **Severe^d^**  **n (%)** |
| --- | --- | --- | --- | --- |
| Any Event^e^ | 369 (38.6) | 295 (30.9) | 217 (22.7) | 75 (7.9) |
| Chills | 710 (74.3) | 143 (15.0) | 72 (7.53) | 31 (3.2) |
| Fatigue | 534 (55.9) | 243 (25.4) | 135 (14.1) | 44 (4.6) |
| Headache | 586 (61.3) | 210 (22.0) | 116 (12.1) | 44 (4.6) |
| Joint pain | 648 (67.8) | 196 (20.5) | 80 (8.4) | 32 (3.4) |
| Malaise | 610 (63.8) | 195 (20.4) | 103 (10.8) | 48 (5.0) |
| Muscle aches | 709 (74.2) | 156 (16.3) | 64 (6.7) | 27 (2.8) |
| Nausea | 785 (82.1) | 114 (11.9) | 35 (3.7) | 22 (2.3) |

^a^ None denotes participants who did not report any systemic reactogenicity event^. e^Any event includes all the systemic reactogenicity events except fever. Participants who reported fever only (8 participants) were considered to have reported no event in this analysis. Fever was not ranked by severity since its severity was not solicited during the follow up period. The severity of the systemic reactogenicity events was determined based on their ability to interfere with the normal daily activities of the participants. ^b^Mild events did not interfere with the normal daily activities of participants. ^c^Moderate events somewhat interfered with the normal daily activities of participants. ^d^Severe events were considerable and prevented the normal daily activities of participants.

**Supplementary Table 8.** Analysis of factors associated with fever

| **Baseline sociodemographic characteristic** | | **Fever** | | **Univariate analysis** | | | **Multivariate analysis^a^** | | |
| --- | --- | --- | --- | --- | --- | --- | --- | --- | --- |
|  | | **n^d^** | **%** | **Odds ratio** | **95% CI** | **p-value^b^** | **Odds ratio** | **95% CI** | **p-value^b^** |
| Age | 17-39yrs. | 158/672 | 23.5 | 1 | 1 | .. | 1 | 1 | .. |
|  | 40-59yrs. | 55/216 | 25.5 | 1.11 | (0.78-1.58) | 0.559 | 0.95 | (0.64-1.40) | 0.792 |
|  | 60+yrs. | 20/68 | 29.4 | 1.36 | (0.78-2.35) | 0.28 | 1.12 | (0.61-2.04) | 0.717 |
| Sex | Male | 58/223 | 26.0 | 1 | 1 | .. | 1 | 1 | .. |
|  | Female, not pregnant | 134/523 | 25.6 | 0.98 | (0.69-1.40) | 0.912 | 1.09 | (0.75-1.59) | 0.648 |
|  | Female, pregnant | 41/210 | 19.5 | 0.69 | (0.44-1.09) | 0.109 | 1.22 | (0.68-2.16) | 0.506 |
| Dose | 1 dose | 126/573 | 22.0 | 1 | 1 | .. | 1 | 1 | .. |
|  | 2 doses, no product mixing^c^ | 27/101 | 27.0 | 1.29 | (0.80-2.10) | 0.295 | 1.36 | (0.83-2.24) | 0.223 |
|  | 2 doses, product mixing^c^ | 32/127 | 25.0 | 1.20 | (0.76-1.87) | 0.434 | 1.24 | (0.76-2.03) | 0.398 |
|  | 3 doses, no product mixing^c^ | 7/30 | 23.0 | 1.08 | (0.45-2.57) | 0.863 | 1.67 | (0.67-4.20) | 0.275 |
|  | 3 doses, product mixing^c^ | 38/116 | 33.0 | 1.73 | (1.12-2.67) | **0.014** | 1.64 | (1.03-2.59) | 0.036 |
|  | 4 doses, product mixing^c^ | 3/9 | 33.0 | 1.77 | (0.44-7.19) | 0.422 | 1.31 | (0.30-5.74) | 0.722 |
| Brand | Pfizer | 62/364 | 17.0 | 1 | 1 | .. | 1 | 1 | .. |
|  | Johnson | 134/492 | 27.2 | 1.82 | (1.30-2.56) | **<0.001** | 2.14 | (1.36-3.36) | **0.001** |
|  | Moderna | 37/100 | 37.0 | 2.86 | (1.75-4.67) | **<0.001** | 2.75 | (1.62-4.68) | **<0.001** |
| Comorbidity | No | 166/691 | 24.0 | 1 | 1 | .. | 1 | 1 | .. |
|  | Yes | 67/265 | 25.3 | 1.07 | (0.77-1.49) | 0.685 | 0.94 | (0.65-1.36) | 0.735 |

Abbreviations: CI, confidence interval; yrs, years. Logistic regression model was used for both univariate and multivariate analysis. ^a^ Multivariate analysis adjusted for all variables in the table. ^b^ P<0.05 was considered statistically significant. ^c^ Product mixing refers to participants who received more than one vaccine brand. The total number of participants was 956. ^d^ n denotes the number of participants who reported fever.

**Supplementary Table 9.** Analysis of factors associated with nausea

| **Baseline sociodemographic characteristic** | | **Nausea** |  | **Univariate analysis** | | | **Multivariate analysis^a^** | | |
| --- | --- | --- | --- | --- | --- | --- | --- | --- | --- |
|  |  | **n^d^** | **%** | **Odds ratio** | **95% CI** | **p-value^b^** | **Odds ratio** | **95% CI** | **p-value^b^** |
| Age | 17-39yrs. | 127/672 | 18.9 | 1 | 1 | .. | 1 | 1 | .. |
|  | 40-59yrs. | 39/216 | 18.1 | 0.95 | (0.64-1.41) | 0.782 | 0.88 | (0.56-1.39) | 0.588 |
|  | 60+yrs. | 5/68 | 7.3 | 0.34 | (0.13-0.86) | **0.023** | 0.38 | (0.14-1.02) | 0.054 |
| Sex | Male | 17/223 | 7.6 | 1 | 1 | .. | 1 | 1 | .. |
|  | Female, not pregnant | 103/523 | 19.7 | 2.97 | (1.73-5.10) | **<0.001** | 2.86 | (1.64-4.98) | **<0.001** |
|  | Female, pregnant | 51/210 | 24.3 | 3.89 | (2.16-6.99) | **<0.001** | 5.62 | (2.75-11.49) | **<0.001** |
| Dose | 1 dose | 101/573 | 17.6 | 1 | 1 | .. | 1 | 1 | .. |
|  | 2 doses, no product mixing^c^ | 17/101 | 16.8 | 0.95 | (0.54-1.66) | 0.846 | 0.92 | (0.51-1.65) | 0.777 |
|  | 2 doses, product mixing^c^ | 21/127 | 16.5 | 0.93 | (0.55-1.55) | 0.769 | 0.87 | (0.49-1.54) | 0.636 |
|  | 3 doses, no product mixing^c^ | 11/30 | 36.7 | 2.71 | (1.25-5.86) | 0.012 | 3.13 | (1.34-7.29) | **0.008** |
|  | 3 doses, product mixing^c^ | 21/116 | 18.1 | 1.03 | (0.62-1.74) | 0.902 | 1.07 | (0.62-1.87) | 0.800 |
|  | 4 doses, product mixing^c^ | 0/9 | 0 | 1 | 1 | .. | 1 | 1 | .. |
| Brand | Pfizer | 66/364 | 18.1 | 1 | 1 | .. | 1 | 1 | .. |
|  | Johnson & Johnson | 81/492 | 16.5 | 0.89 | (0.62-1.27) | 0.522 | 1.73 | (1.03-2.9) | **0.037** |
|  | Moderna | 24/100 | 24 | 1.43 | (0.84-2.42) | 0.190 | 2.31 | (1.28-4.2) | **0.006** |
| Comorbidity | No | 116/691 | 16.8 | 1 | 1 | .. | 1 | 1 | .. |
|  | Yes | 55/265 | 20.8 | 1.30 | (0.91-1.86) | 0.153 | 1.68 | (1.10-2.57) | **0.017** |

Abbreviations: CI, confidence interval; yrs, years. Logistic regression model was used for both univariate and multivariate analysis. ^a^ Multivariate analysis adjusted for all variables in the table. ^b^ P<0.05 was considered statistically significant. ^c^ Product mixing refers to participants who received more than one vaccine brand. The total number of participants was 956. ^d^ n denotes the number of participants who reported nausea.

**Supplementary Table 10.** Analysis of factors associated with malaise.

| **Baseline sociodemographic characteristic** | | **Malaise** | | **Univariate analysis** | | | **Multivariate analysis^a^** | | |
| --- | --- | --- | --- | --- | --- | --- | --- | --- | --- |
|  | | **n^d^** | **%** | **Odds ratio** | **95% CI** | **p-value^b^** | **Odds ratio** | **95% CI** | **p-value^b^** |
| Age | 17-39yrs. | 250/672 | 37.2 | 1 | 1 | .. | 1 | 1 | .. |
|  | 40-59yrs. | 79/216 | 36.6 | 0.97 | (0.71-1.34) | 0.868 | 0.76 | (0.53-1.09) | 0.134 |
|  | 60+yrs. | 17/68 | 25.0 | 0.56 | (0.32-1.00) | **0.048** | 0.47 | (0.25-0.86) | **0.015** |
| Sex | Male | 69/223 | 30.9 | 1 | 1 | .. | 1 | 1 | .. |
|  | Female, not pregnant | 210/523 | 40.2 | 1.50 | (1.07-2.09) | **0.018** | 1.57 | (1.10-2.23) | 0.012 |
|  | Female, pregnant | 67/210 | 31.9 | 1.05 | (0.70-1.57) | 0.829 | 1.55 | (0.92-2.59) | 0.098 |
| Dose | 1 dose | 205/573 | 35.8 | 1 | 1 | .. | 1 | 1 | .. |
|  | 2 doses, no product mixing^c^ | 34/101 | 33.7 | 0.91 | (0.58-1.42) | 0.682 | 0.92 | (0.58-1.46) | 0.721 |
|  | 2 doses, product mixing^c^ | 49/127 | 38.6 | 1.13 | (0.76-1.68) | 0.552 | 1.08 | (0.70-1.68) | 0.725 |
|  | 3 doses, no product mixing^c^ | 12/30 | 40.0 | 1.20 | (0.57-2.53) | 0.639 | 1.56 | (0.71-3.47) | 0.272 |
|  | 3 doses, product mixing^c^ | 44/116 | 37.9 | 1.10 | (0.73-1.66) | 0.660 | 1.03 | (0.66-1.59) | 0.911 |
|  | 4 doses, product mixing^c^ | 2/9 | 22.2 | 0.51 | (0.11-2.49) | 0.408 | 0.60 | (0.12-3.12) | 0.542 |
| Brand | Pfizer | 104/364 | 28.6 | 1 | 1 | .. | 1 | 1 | .. |
|  | Johnson & Johnson | 191/492 | 38.8 | 1.59 | (1.19-2.12) | **0.002** | 1.99 | (1.34-2.96) | **0.001** |
|  | Moderna | 51/100 | 51.0 | 2.60 | (1.65-4.09) | **<0.001** | 3.06 | (1.87-5.02) | **<0.001** |
| Comorbidity | No | 241/691 | 34.9 | 1 | 1 | .. | 1 | 1 | .. |
|  | Yes | 105/265 | 39.6 | 1.23 | (0.92-1.64) | 0.172 | 1.30 | (0.93-1.82) | 0.127 |

Abbreviations: CI, confidence interval; yrs, years. Logistic regression model was used for both univariate and multivariate analysis. ^a^ Multivariate analysis adjusted for all variables in the table. ^b^ P<0.05 was considered statistically significant. ^c^ Product mixing refers to participants who received more than one vaccine brand. The total number of participants was 956. ^d^ n denotes the number of participants who reported malaise.

**Supplementary Table 11.** Analysis of factors associated with chills.

| **Baseline sociodemographic characteristic** | | **Chills** | | **Univariate analysis** | | | **Multivariate analysis^a^** | | |
| --- | --- | --- | --- | --- | --- | --- | --- | --- | --- |
|  | | **n^d^** | **%** | **Odds ratio** | **95% CI** | **p-value^b^** | **Odds ratio** | **95% CI** | **p-value^b^** |
| Age | 17-39yrs. | 177/672 | 26.3 | 1 | 1 | .. | 1 | 1 | .. |
|  | 40-59yrs. | 57/216 | 26.4 | 1.00 | (0.71-1.42) | 0.989 | 0.78 | (0.53-1.15) | 0.214 |
|  | 60+yrs. | 12/68 | 17.6 | 0.60 | (0.31-1.14) | 0.121 | 0.53 | (0.26-1.05) | 0.069 |
| Sex | Male | 46/223 | 20.6 | 1 | 1 | .. | 1 | 1 | .. |
|  | Female, not pregnant | 163/523 | 31.2 | 1.74 | (1.20-2.53) | **0.004** | 1.86 | (1.26-2.76) | **0.002** |
|  | Female, pregnant | 37/210 | 17.6 | 0.82 | (0.51-1.33) | 0.427 | 1.30 | (0.72-2.36) | 0.382 |
| Dose | 1 dose | 138/573 | 24.1 | 1 | 1 | **..** | 1 | 1 | **..** |
|  | 2 doses, no product mixing^c^ | 18/101 | 17.8 | 0.68 | (0.40-1.18) | 0.171 | 0.68 | (0.39-1.20) | 0.183 |
|  | 2 doses, product mixing^c^ | 35/127 | 27.6 | 1.20 | (0.78-1.85) | 0.412 | 1.11 | (0.68-1.83) | 0.668 |
|  | 3 doses, no product mixing^c^ | 13/30 | 43.3 | 2.41 | (1.14-5.09) | **0.021** | 3.48 | (1.54-7.88) | **0.003** |
|  | 3 doses, product mixing^c^ | 41/116 | 35.3 | 1.72 | (1.13-2.64) | **0.012** | 1.53 | (0.97-2.43) | 0.069 |
|  | 4 doses, product mixing^c^ | 1/9 | 11.1 | 0.39 | (0.05-3.18) | 0.382 | 0.37 | (0.04-3.19) | 0.365 |
| Brand | Pfizer | 63/364 | 17.3 | 1 | 1 | .. | 1 | 1 | .. |
|  | Johnson & Johnson | 139/492 | 28.3 | 1.88 | (1.35-2.63) | **<0.001** | 2.41 | (1.53-3.78) | **<0.001** |
|  | Moderna | 44/100 | 44.0 | 3.75 | (2.33-6.06) | **<0.001** | 4.11 | (2.42-6.98) | **<0.001** |
| Comorbidity | No | 175/691 | 25.3 | 1 | 1 | .. | 1 | 1 | .. |
|  | Yes | 71/265 | 26.8 | 1.08 | (0.78-1.49) | 0.642 | 1.04 | (0.72-1.52) | 0.82 |

Abbreviations: CI, confidence interval; yrs, years. Logistic regression model was used for both univariate and multivariate analysis. ^a^ Multivariate analysis adjusted for all variables in the table. ^b^ P<0.05 was considered statistically significant. ^c^ Product mixing refers to participants who received more than one vaccine brand. The total number of participants was 956. ^d^ n denotes the number of participants who reported chills.

**Supplementary Table 12.** Analysis of factors associated with headache.

| **Baseline sociodemographic characteristic** | | **Headache** |  | **Univariate analysis** | | | **Multivariate analysis^a^** | | |
| --- | --- | --- | --- | --- | --- | --- | --- | --- | --- |
|  | | **n^d^** | **%** | **Odds ratio** | **95% CI** | **p-value^b^** | **Odds ratio** | **95% CI** | **p-value^b^** |
| Age | 17-39yrs. | 262/672 | 39.0 | 1 | 1 | .. | 1 | 1 | .. |
|  | 40-59yrs. | 88/216 | 40.7 | 1.08 | (0.79-1.47) | 0.647 | 0.87 | (0.61-1.24) | 0.443 |
|  | 60+yrs. | 20/68 | 29.4 | 0.65 | (0.39-1.12) | 0.123 | 0.52 | (0.29-0.94) | **0.030** |
| Sex | Male | 69/223 | 30.9 | 1 | 1 | .. | 1 | 1 | .. |
|  | Female, not pregnant | 230/523 | 44.0 | 1.75 | (1.26-2.44) | **0.001** | 1.79 | (1.26-2.54) | **0.001** |
|  | Female, pregnant | 71/210 | 33.8 | 1.14 | (0.76-1.71) | 0.524 | 1.53 | (0.92-2.55) | 0.099 |
| Dose | 1 dose | 212/573 | 37.0 | 1 | 1 | .. | 1 | 1 | .. |
|  | 2 doses, no product mixing^c^ | 34/101 | 33.7 | 0.86 | (0.55-1.35) | 0.521 | 0.82 | (0.52-1.30) | 0.405 |
|  | 2 doses, product mixing^c^ | 49/127 | 38.6 | 1.07 | (0.72-1.59) | 0.738 | 0.96 | (0.62-1.48) | 0.837 |
|  | 3 doses, no product mixing^c^ | 15/30 | 50.0 | 1.70 | (0.82-3.55) | 0.156 | 1.86 | (0.85-4.04) | 0.119 |
|  | 3 doses, product mixing^c^ | 56/116 | 48.3 | 1.59 | (1.06-2.38) | **0.024** | 1.48 | (0.97-2.27) | 0.072 |
|  | 4 doses, product mixing^c^ | 4/9 | 44.4 | 1.36 | (0.36-5.13) | 0.648 | 1.70 | (0.41-6.96) | 0.464 |
| Brand | Pfizer | 121/364 | 33.2 | 1 | 1 | .. | 1 | 1 | .. |
|  | Johnson & Johnson | 195/492 | 39.6 | 1.32 | (0.99-1.75) | 0.056 | 1.50 | (1.02-2.19) | **0.040** |
|  | Moderna | 54/100 | 54.0 | 2.36 | (1.50-3.70) | **<0.001** | 2.49 | (1.53-4.06) | **<0.001** |
| Comorbidity | No | 255/691 | 36.9 | 1 | 1 | .. | 1 | 1 | .. |
|  | Yes | 115/265 | 43.4 | 1.31 | (0.98-1.75) | 0.065 | 1.40 | (1.00-1.96) | **0.048** |

Abbreviations: CI, confidence interval; yrs, years. Logistic regression model was used for both univariate and multivariate analysis. ^a^ Multivariate analysis adjusted for all variables in the table. ^b^ P<0.05 was considered statistically significant. ^c^ Product mixing refers to participants who received more than one vaccine brand. The total number of participants was 956. ^d^ n denotes the number of participants who reported headache.

**Supplementary Table 13.** Analysis of factors associated with joint pain.

| **Baseline sociodemographic characteristic** | | **Joint pain** |  | **Univariate analysis** | | | **Multivariate analysis^a^** | | |
| --- | --- | --- | --- | --- | --- | --- | --- | --- | --- |
|  | | **n^d^** | **%** | **Odds ratio** | **95% CI** | **p-value^b^** | **Odds ratio** | **95% CI** | **p-value^b^** |
| Age | 17-39yrs. | 221/672 | 32.9 | 1 | 1 | .. | 1 | 1 | .. |
|  | 40-59yrs. | 68/216 | 31.5 | 0.94 | (0.68-1.30) | 0.701 | 0.76 | (0.52-1.09) | 0.138 |
|  | 60+yrs. | 19/68 | 27.9 | 0.79 | (0.46-1.38) | 0.407 | 0.71 | (0.39-1.28) | 0.253 |
| Sex | Male | 66/223 | 29.6 | 1 | 1 | .. | 1 | 1 | .. |
|  | Female, not pregnant | 180/523 | 34.4 | 1.25 | (0.89-1.75) | 0.200 | 1.35 | (0.94-1.93) | 0.100 |
|  | Female, pregnant | 62/210 | 29.5 | 1.00 | (0.66-1.51) | 0.987 | 1.65 | (0.97-2.81) | 0.066 |
| Dose | 1 dose | 181/573 | 31.6 | 1 | 1 | .. | 1 | 1 | .. |
|  | 2 doses, no product mixing^c^ | 26/101 | 25.7 | 0.75 | (0.47-1.21) | 0.241 | 0.77 | (0.47-1.25) | 0.287 |
|  | 2 doses, product mixing^c^ | 46/127 | 36.2 | 1.23 | (0.82-1.84) | 0.313 | 1.17 | (0.74-1.84) | 0.497 |
|  | 3 doses, no product mixing^c^ | 9/30 | 30.0 | 0.93 | (0.42-2.07) | 0.855 | 1.35 | (0.58-3.17) | 0.489 |
|  | 3 doses, product mixing^c^ | 45/116 | 38.8 | 1.37 | (0.91-2.07) | 0.133 | 1.27 | (0.82-1.97) | 0.288 |
|  | 4 doses, product mixing^c^ | 1/9 | 11.1 | 0.27 | (0.03-2.18) | 0.220 | 0.23 | (0.03-1.98) | 0.181 |
| Brand | Pfizer | 87/364 | 23.9 | 1 | 1 | .. | 1 | 1 | .. |
|  | Johnson & Johnson | 170/492 | 34.5 | 1.68 | (1.24-2.28) | **0.001** | 2.21 | (1.46-3.36) | **<0.001** |
|  | Moderna | 51/100 | 51.0 | 3.31 | (2.09-5.25) | **<0.001** | 3.80 | (2.30-6.30) | **<0.001** |
| Comorbidity | No | 216/691 | 31.3 | 1 | 1 | .. | 1 | 1 | .. |
|  | Yes | 92/265 | 34.7 | 1.17 | (0.87-1.58) | 0.306 | 1.24 | (0.88-1.75) | 0.227 |

Abbreviations: CI, confidence interval; yrs, years. Logistic regression model was used for both univariate and multivariate analysis. ^a^ Multivariate analysis adjusted for all variables in the table. ^b^ P<0.05 was considered statistically significant. ^c^ Product mixing refers to participants who received more than one vaccine brand. The total number of participants was 956. ^d^ n denotes the number of participants who reported joint pain.

**Supplementary Table 14.** Analysis of factors associated with muscle aches.

| **Baseline sociodemographic characteristic** | | **Muscle aches** |  | **Univariate analysis** | | | **Multivariate analysis^a^** | | |
| --- | --- | --- | --- | --- | --- | --- | --- | --- | --- |
|  | | **n^d^** | **%** | **Odds ratio** | **95% CI** | **p-value^b^** | **Odds ratio** | **95% CI** | **p-value^b^** |
| Age | 17-39yrs. | 170/672 | 25.3 | 1 | 1 | .. | 1 | 1 | .. |
|  | 40-59yrs. | 59/216 | 27.3 | 1.11 | (0.79-1.57) | 0.556 | 0.83 | (0.57-1.22) | 0.352 |
|  | 60+yrs. | 18/68 | 26.5 | 1.06 | (0.60-1.87) | 0.832 | 0.81 | (0.44-1.49) | 0.494 |
| Sex | Male | 60/223 | 26.9 | 1 | 1 | .. | 1 | 1 | .. |
|  | Female, not pregnant | 156/523 | 29.8 | 1.16 | (0.81-1.64) | 0.421 | 1.20 | (0.83-1.74) | 0.324 |
|  | Female, pregnant | 31/210 | 14.8 | 0.47 | (0.29-0.76) | **0.002** | 0.67 | (0.38-1.21) | 0.188 |
| Dose | 1 dose | 134/573 | 23.4 | 1 | 1 | .. | 1 | 1 | .. |
|  | 2 doses, no product mixing^c^ | 24/101 | 23.8 | 1.02 | (0.62-1.68) | 0.934 | 1.01 | (0.61-1.69) | 0.958 |
|  | 2 doses, product mixing^c^ | 40/127 | 31.5 | 1.51 | (0.99-2.30) | 0.057 | 1.44 | (0.89-2.32) | 0.134 |
|  | 3 doses, no product mixing^c^ | 9/30 | 30.0 | 1.40 | (0.63-3.14) | 0.408 | 1.95 | (0.82-4.65) | 0.132 |
|  | 3 doses, product mixing^c^ | 38/116 | 32.8 | 1.60 | (1.04-2.46) | **0.034** | 1.36 | (0.86-2.16) | 0.193 |
|  | 4 doses, product mixing^c^ | 2/9 | 22.2 | 0.94 | (0.19-4.56) | 0.935 | 0.66 | (0.13-3.46) | 0.622 |
| Brand | Pfizer | 62/364 | 17.0 | 1 | 1 | .. | 1 | 1 | .. |
|  | Johnson & Johnson | 143/492 | 29.1 | 2.00 | (1.43-2.79) | **<0.001** | 1.89 | (1.22-2.92) | **0.004** |
|  | Moderna | 42/100 | 42.0 | 3.53 | (2.18-5.71) | **<0.001** | 3.11 | (1.85-5.23) | **<0.001** |
| Comorbidity | No | 171/691 | 24.8 | 1 | 1 | .. | 1 | 1 | .. |
|  | Yes | 76/265 | 28.7 | 1.22 | (0.89-1.68) | 0.214 | 1.08 | (0.75-1.56) | 0.664 |

Abbreviations: CI, confidence interval; yrs, years. Logistic regression model was used for both univariate and multivariate analysis. ^a^ Multivariate analysis adjusted for all variables in the table. ^b^ P<0.05 was considered statistically significant. ^c^ Product mixing refers to participants who received more than one vaccine brand. The total number of participants was 956. ^d^ n denotes the number of participants who reported muscle aches.

**Supplementary Table 15.** Analysis of factors associated with fatigue.

| **Baseline sociodemographic characteristic** | | **Fatigue** |  | **Univariate analysis** | | | **Multivariate analysis^a^** | | |
| --- | --- | --- | --- | --- | --- | --- | --- | --- | --- |
|  | | **n^d^** | **%** | **Odds ratio** | **95% CI** | **p-value^b^** | **Odds ratio** | **95% CI** | **p-value^b^** |
| Age | 17-39yrs. | 302/672 | 44.9 | 1 | 1 | .. | 1 | 1 | .. |
|  | 40-59yrs. | 97/216 | 44.9 | 1.00 | (0.73-1.36) | 0.993 | 0.83 | (0.59-1.18) | 0.307 |
|  | 60+yrs. | 23/68 | 33.8 | 0.63 | (0.37-1.06) | **0.080** | 0.52 | (0.29-0.93) | **0.027** |
| Sex | Male | 87/223 | 39.0 | 1 | 1 | .. | 1 | 1 | .. |
|  | Female, not pregnant | 246/523 | 47.0 | 1.39 | (1.01-1.91) | **0.044** | 1.55 | (1.11-2.18) | **0.011** |
|  | Female, pregnant | 89/210 | 42.4 | 1.15 | (0.78-1.69) | 0.476 | 2.12 | (1.28-3.51) | **0.003** |
| Dose | 1 dose | 242/573 | 42.2 | 1 | 1 | .. | 1 | 1 | .. |
|  | 2 doses, no product mixing^c^ | 42/101 | 41.6 | 0.97 | (0.63-1.50) | 0.903 | 1.04 | (0.67-1.62) | 0.867 |
|  | 2 doses, product mixing^c^ | 63/127 | 49.6 | 1.35 | (0.92-1.98) | 0.130 | 1.33 | (0.86-2.05) | 0.199 |
|  | 3 doses, no product mixing^c^ | 13/30 | 43.3 | 1.05 | (0.50-2.19) | 0.905 | 1.57 | (0.71-3.47) | 0.261 |
|  | 3 doses, product mixing^c^ | 58/116 | 50.0 | 1.37 | (0.92-2.04) | 0.125 | 1.31 | (0.85-2.02) | 0.216 |
|  | 4 doses, product mixing^c^ | 4/9 | 44.4 | 1.09 | (0.29-4.12) | 0.894 | 1.11 | (0.27-4.64) | 0.882 |
| Brand | Pfizer | 125/364 | 34.3 | 1 | 1 | .. | 1 | 1 | .. |
|  | Johnson & Johnson | 229/492 | 46.5 | 1.67 | (1.26-2.20) | **<0.001** | 2.53 | (1.71-3.74) | **<0.001** |
|  | Moderna | 68/100 | 68.0 | 4.06 | (2.53-6.52) | **<0.001** | 4.75 | (2.83-7.96) | **<0.001** |
| Comorbidity | No | 302/691 | 43.7 | 1 | 1 | .. | 1 | 1 | .. |
|  | Yes | 120/265 | 45.3 | 1.07 | (0.80-1.42) | 0.660 | 1.11 | (0.80-1.55) | 0.526 |

Abbreviations: CI, confidence interval; yrs, years. Logistic regression model was used for both univariate and multivariate analysis. ^a^ Multivariate analysis adjusted for all variables in the table. ^b^ P<0.05 was considered statistically significant. ^c^ Product mixing refers to participants who received more than one vaccine brand. The total number of participants was 956. ^d^ n denotes the number of participants who reported fatigue.

**Supplementary Table 16.** Summary of pregnancy related post-vaccination hospitalization events within the cohort.

|  | **Age in years** | **Reported event** | **Time of event onset in days relative to the date of vaccination** | **Vaccine name** | **Vaccine dose** |
| --- | --- | --- | --- | --- | --- |
| 1. | 38 | Non-reassuring fetal status | 11 | Pfizer | 1^st^ Vaccination |
| 2. | 33 | Incomplete abortion | 16 | Pfizer | 3^rd^ Vaccination |
| 3. | 28 | Incomplete abortion | 22 | Pfizer | 3^rd^ Vaccination |
| 4. | 24 | High blood pressure in pregnancy | 34 | Moderna | 2^nd^ Vaccination |
| 5. | 22 | Antepartum haemorrhage | 39 | Pfizer | 1^st^ Vaccination |
| 6. | 29 | Spontaneous abortion | 40 | Moderna | 2^nd^ Vaccination |
| 7. | 33 | Incomplete abortion | 45 | Pfizer | 2^nd^ Vaccination |
| 8. | 24 | High blood pressure in pregnancy | 55 | Moderna | 1^st^ Vaccination |
| 9. | 23 | Premature delivery | 57 | Pfizer | 1^st^ Vaccination |
| 10. | 27 | Threatened abortion | 66 | Pfizer | 2^nd^ Vaccination |
| 11. | 27 | Spontaneous abortion | 84 | Pfizer | 1^st^ Vaccination |
| 12. | 19 | Antepartum haemorrhage | 84 | Pfizer | 1^st^ Vaccination |
| 13. | 21 | Complicated pregnancy | 84 | Pfizer | 1^st^ Vaccination |
| 14. | 26 | Preterm premature rupture of membranes (PPROM) at 33 weeks of pregnancy | 88 | Moderna | 1^st^ Vaccination |

**Supplementary Table 17.** Summary of elective surgeries reported as post-vaccination hospitalization events within the cohort.

|  | **Age** | **Reported event** | **Time of event onset in days relative to the date of vaccination** | **Vaccine name** | **Vaccine dose** |
| --- | --- | --- | --- | --- | --- |
| 1. | 68 | Inguinal hernia | 34 | Moderna | 2^nd^ Vaccination |
| 2. | 61 | Lipoma (Armpit region) | 47 | Johnson & Johnson | 2^nd^ Vaccination |
| 3. | 60 | Intestinal obstruction | 89 | Johnson & Johnson | 2^nd^ Vaccination |

**Supplementary Table 18.** Summary of other medical conditions reported as post-vaccination hospitalization events within the cohort.

|  | **Age in years** | **Reported event(s)** | **Time of event onset in days relative to the date of vaccination** | **Vaccine name** | **Vaccine dose** |
| --- | --- | --- | --- | --- | --- |
| 1. | 32 | Dengue fever | 22 | Johnson & Johnson | 1^st^ Vaccination |
| 2. | 36 | High blood pressure | 27 | Pfizer | 2^nd^ Vaccination |
| 3. | 64 | Vomiting | 34 | Johnson & Johnson | 1^st^ Vaccination |
| 4. | 25 | Generalized malaise | 41 | Pfizer | 1^st^ Vaccination |
| 5. | 23 | Malaria | 44 | Johnson & Johnson | 1^st^ Vaccination |
| 6. | 26 | Dehydration | 49 | Johnson & Johnson | 3^rd^ Vaccination |
| 7. | 64 | Chronic kidney failure and cancer | 55 | Johnson & Johnson | 1^st^ Vaccination |
| 8. | 56 | Diarrhoea | 55 | Johnson & Johnson | 1^st^ Vaccination |
| 9. | 20 | Headache, malaise, and weakness | 89 | Pfizer | 1^st^ Vaccination |
| 10. | 20 | Headache, fever, and abdominal pain | 71 | Pfizer | 1^st^ Vaccination |
| 11. | 56 | Pneumonia | 77 | Moderna | 2^nd^ Vaccination |
| 12. | 47 | Cervical cancer | 80 | Johnson & Johnson | 1^st^ Vaccination |
| 13. | 21 | Malaria | 81 | Pfizer | 1^st^ Vaccination |
| 14. | 39 | Lung bulge | 105 | Pfizer | 1^st^ Vaccination |

**Supplementary Table 19.** Non-hospitalization events reported by study participants

|  | **Age in years** | **Reported event (s)** | **Time of event onset in days relative to the date of vaccination** | **Vaccine name** | **Vaccine dose** |
| --- | --- | --- | --- | --- | --- |
| 1 | 25 | Joint pains, dizziness, and stoppage of menses | 0 | Johnson & Johnson | 1^st^ Booster |
| 2 | 50 | Headache and heart palpitations | 2 | Pfizer | 1^st^ Booster |
| 3 | 40 | Reduced breast milk production, and blurred vision | 3 | Johnson & Johnson | 1^st^ Vaccination |
| 4 | 25 | Left breast swelling | 4 | Johnson & Johnson | 1^st^ Vaccination |
| 5 | 34 | Itchiness of left foot followed by blister | 4 | Johnson & Johnson | 1st Vaccination |
| 6 | 53 | Itchiness and numbness of left lower limb | 52 | Johnson & Johnson | 1^st^ Vaccination |

**Supplementary Table 20.** Analysis of SMS response rate among a subset of non-reactogenicity participants.

| **Week of follow-up** | **Participants targeted^a^**  **N=90** | **No. of participants with complete response^b^**  **n (%)** | **No. of participants with partial response^c^**  **n (%)** | **No. of participants who didn’t respond^d^**  **n (%)** | **No. of participants who responded incorrectly^e^**  **n (%)** |
| --- | --- | --- | --- | --- | --- |
| 1 | 16 | 8 (50.0) | 0 (0.0) | 0 (0.0) | 8 (50.0) |
| 2 | 20 | 8 (40.0) | 1 (5.0) | 0 (0.0) | 11 (55.0) |
| 3 | 20 | 10 (50.0) | 0 (0.0) | 0 (0.0) | 10 (50.0) |
| 4 | 36 | 10 (27.8) | 0 (0.0) | 4 (11.1) | 22 (61.1) |
| 5 | 73 | 25 (34.2) | 0 (0.0) | 10 (13.7) | 38 (52.1) |
| 6 | 94 | 26 (27.7) | 2 (2.1) | 10 (10.6) | 56 (59.6) |
| 7 | 91 | 29 (31.9) | 0 (0.0) | 10 (11.0) | 52 (57.1) |
| 8 | 89 | 32 (36.0) | 2 (2.2) | 13 (14.6) | 42 (47.2) |
| 9 | 87 | 29 (33.3) | 2 (2.3) | 31 (35.6) | 25 (28.7) |
| 10 | 83 | 32 (38.6) | 1 (1.2) | 43 (51.8) | 7 (8.4) |
| 11 | 69 | 27 (39.1) | 5 (7.2) | 33 (47.8) | 4 (5.8) |
| 12 | 68 | 23 (33.8) | 0 (0.0) | 41 (60.3) | 4 (5.9) |
| 13 | 68 | 21 (30.9) | 0 (0.0) | 41 (60.3) | 6 (8.8) |
| Total | 814 | 280 (34.4) | 13 (1.6) | 236 (29.0) | 285 (35.0) |

Abbreviations: SMS, Short Message Service; No., Number. A total of 90 non-reactogenicity participants were followed up using the SMS platform during their weekly follow-up (Week 1-13) to gather information on post-vaccination events. ^a^ Participants targeted denotes participants due for follow-up by SMS for that week. ^b^ No. of participants with complete response denotes participants who responded correctly to all the SMS sent to them. ^c^ No. of participants with partial response denotes participants who responded only to some of the SMS sent to them. ^d^ No. of participants who didn’t respond denotes participants who did not answer any of the SMS sent to them. ^e^ No. of participants who responded incorrectly denotes participants who provided invalid responses (wrong answers) to the SMS sent to them.
