## Supplementary material for "Active vaccine safety surveillance: Experience from a prospective cohort event monitoring study of COVID-19 vaccines in Kenya": Data collection tools-Study questionnaires

**CEM for COVID-19 vaccine safety**

***Baseline questionnaire: Participant registration, informed consent, contact, and covariates***

| Question | Response value/ coding or type |
| --- | --- |
| 1. Unique identifier for each site | Unique Numeric/ text/ alphanumeric |
| 2. Unique identifier for each participant | Unique Numeric/ text/ alphanumeric |
| 3. Name of participant | |
| 3.1 First name | Text |
| 3.2 Middle name | Text |
| 3.3 Surname | Text |
| 4. Informed consent provided? | 0 = No<br>1 = Yes |
| 5. Is the participant part of the reactogenicity subset | 0 = No<br>1 = Yes |
| 6. Participant contact details | |
| 6.1. Participant's phone number | Numeric |
| 6.2. Name of next of kin | Text |
| 6.3. Phone number of next of kin | Numeric |
| 7. Participant covariates | |
| 7.1. Date of birth | dd/mm/yyyy |
| 7.2. Sex | 0 = Male<br>1 = Female<br>2 = Other |
| 7.3. Is the participant pregnant? | 0 = No<br>1 = Yes |
| 7.4. Is the participant breastfeeding? | 0 = No<br>1 = Yes |
| 7.5. Medical history, presence of diseases? | 0 = No medical history<br>1 = Chronic respiratory disease or asthma<br>2 = Chronic heart disease<br>3 = Chronic liver disease<br>4 = Chronic renal disease<br>5 = Diabetes<br>6 = Immunocompromised/ immunosuppressed<br>7 = Obesity<br>8 = Allergy |
| 7.6. Previous COVID-19 disease? | 0 = No<br>1 = Yes, laboratory confirmed<br>2 = Probable but not laboratory-confirmed |
| 7.7. History of reaction to any vaccination? | 0 = No<br>1 = Yes<br>2 = Do not know |

| Question | Response value/ coding or type |
|---|---|
| 7.7.1. **If yes**: Indicate which vaccine | 1= BCG (Tuberculosis vaccine)<br>2= Polio vaccine<br>3=DPT (diphtheria, pertussis & tetanus) vaccine<br>4=Pentavalent vaccine (diphtheria, pertussis, tetanus, hepatitis B and haemophilus influenzae type B vaccines)<br>5=Hepatitis B vaccine<br>6=Hemophilus influenzae type B vaccine<br>7= MMR (measles, mumps, and rubella vaccine)<br>8=Yellow fever vaccine<br>9=Tetanus toxoid vaccine<br>10=HPV (human papillomavirus) vaccine<br>11=Hepatitis A vaccine<br>12=Hepatitis C vaccine<br>13=Typhoid fever vaccine<br>14=Rabies vaccine.<br>15=Pneumococcal conjugate vaccine<br>16=Cholera vaccine<br>17=Varicella vaccine<br>18=COVID-19 vaccine<br>19= Meningococcal vaccine<br>20=Others(specify) |

| Question | Response value/ coding or type |
|---|---|
| 7.7.2. **Describe the reaction to vaccine** | 1=Pain at injection site<br>2=Redness around injection site<br>3=swelling around injection site<br>4=bruise(haematoma) around injection site<br>5=Itching around injection site<br>6=warmth around injection site<br>7=Fever (hotness of body)<br>8=Chills (feeling of being cold)<br>9=Headache<br>10=nausea (feeling of wanting to vomit)<br>11=vomiting<br>12=Muscle ache<br>13=Joint pain<br>14=Malaise (feeling of weakness/not well)<br>15=fatigue (feeling tired)<br>16=urticaria (hives/weal/allergic rashes)<br>17= Anaphylactic shock<br>18=other (Specify) |
| 8. COVID-19 vaccine exposure information | |
| 8.1. Dose of **current** COVID-19 vaccine | 1 = 1st vaccination<br>2 = 2nd vaccination<br>3 = 3rd vaccination<br>4 = 4th vaccination<br>5 = other (specify) |
| 8.2. Date of **current** vaccination | dd/mm/yyyy |
| 8.3. Time of **current** vaccination | HH:MM |
| 8.4. Vaccine brand/ manufacturer | 1= AstraZeneca<br>2= Pfizer<br>3= Johnson & Johnson<br>4= Moderna<br>5= Sputnik V<br>6= Sinopharm<br>7= Other (specify<br>8= Don’t know |
| 8.5. Vaccine batch number | Text |
| 8.6. **Pre-populated based on response to 8.4**: Was a separate diluent required? | 0 = No<br>1 = Yes |
| 8.6.1. **If yes**: Diluent brand/ manufacturer | 1= AstraZeneca<br>2= Pfizer<br>3= Johnson & Johnson<br>4= Moderna<br>5= Sputnik V<br>6= Sinopharm<br>7= Other (specify<br>8= Don’t know |

| Question | Response value/ coding or type |
|---|---|
| 8.6.2. Diluent batch number | Text |
| 8.7. Co-administration of vaccine against any other disease other than COVID | 0 = No<br>1 = Yes |
| 8.7.1. **If yes**: Specify which disease was vaccinated against | Text |
| 8.8. Received previous dose of COVID vaccine? | 0 = No<br>1 = Yes |
| 8.8.1. **If yes**: Number of previous doses | Text |
| 8.8.2. Dose 1 confirmed using: | 0 = Verbal report<br>1 = SMS<br>2 = Vaccination certificate |
| 8.8.2.1. Dose 1 date | dd/mm/yyyy |
| 8.8.2.2. Dose 1 vaccine brand/ manufacturer | 1= AstraZeneca<br>2= Pfizer<br>3= Johnson & Johnson<br>4= Moderna<br>5= Sputnik V<br>6= Sinopharm<br>7= Other (specify<br>8= Don't know |
| 8.8.3. Dose 2 confirmed using: | 0 = Verbal report<br>1 = SMS<br>2 = Vaccination certificate |
| 8.8.3.1. Dose 2 date | dd/mm/yyyy |
| 8.8.3.2. Dose 2 vacc/ine brand/ manufacturer | 1= AstraZeneca<br>2= Pfizer<br>3= Johnson & Johnson<br>4= Moderna<br>5= Sputnik V<br>6= Sinopharm<br>7= Other (specify<br>8= Don't know |
| 8.8.4. Dose 3 confirmed using: | 0 = Verbal report<br>1 = SMS<br>2 = Vaccination certificate |
| 8.8.4.1. Dose 3 date | dd/mm/yyyy |
| 8.8.4.2. Dose 3 vaccine brand/ manufacturer | 1= AstraZeneca<br>2= Pfizer<br>3= Johnson & Johnson<br>4= Moderna<br>5= Sputnik V<br>6= Sinopharm<br>7= Other (specify<br>8= Don't know |

***Pre-vaccine reactogenicity questionnaire, reactogenicity subset (if the response to Q5 is 'yes')***

| Question | Response value/ coding or type |
|---|---|
| 9. **If Q5 is yes:** Pre-vaccine reactogenicity | |
| 9.1. Unique ID for the record | Unique Numeric/ text/ alphanumeric |
| 9.2. Date of record | dd/mm/yyyy |
| 9.3. Did you feel **feverish** in the past 3 days? | 0 = No<br>1 = Yes |
| 9.3.1. **If yes:** Did you measure your temperature in the past 3 days? | 0 = No<br>1 = Yes |
| 9.3.2. **If yes:** What was your temperature? | 1 = Below 38.0°C (below 100.4°F)<br>2 = 38.0°C to 38.4°C (100.4°F to 101.12 °F)<br>3 = 38.5°C to 38.9°C (101.3°F to 102.02°F)<br>4 = Higher than 39.0°C (higher than 102.2°F)<br>5= I can't remember |
| 9.3.3. How did you measure the temperature? | 1 = Oral (in the mouth)<br>2 = Rectum (in the anus)<br>3 = Armpit<br>4 = Ear<br>5 = Forehead |
| 9.4. Did you feel **like vomiting (nausea)** in the past 3 days, or did you vomit? | 0 = No<br>1 = Yes |
| 9.4.1. **If yes:** How severe was the nausea/ vomiting? | 1 = The nausea/vomiting did not interfere with my activities<br>2 = The nausea/vomiting somewhat interfered with my activities<br>3 = The nausea/vomiting was considerable and prevented my daily activities |
| 9.5. Did you experience **a feeling or weakness or not feeling well (malaise)** in the past 3 days? | 0 = No<br>1 = Yes |
| 9.5.1. **If yes:** How severe was the malaise? | 1 = The malaise did not interfere with my activities<br>2 = The malaise somewhat interfered with my activities<br>3 = The malaise was considerable and prevented my daily activities |
| 9.6. Did you have **a feeling of being cold (chills)** in the past 3 days? | 0 = No<br>1 = Yes |
| 9.6.1. **If yes:** How severe were the chills? | 1 = The chills did not interfere with my activities<br>2 = The chills somewhat interfered with my activities<br>3 = The chills were considerable and prevented my daily activities |
| 9.7. Did you have a **headache** in the past 3 days? | 0 = No<br>1 = Yes |

| Question | Response value/ coding or type |
|---|---|
| 9.7.1. **If yes:** How bad was the headache? | 1 = The headache did not interfere with my activities<br>2 = The headache somewhat interfered with my activities<br>3 = The headache was considerable and prevented my daily activities |
| 9.8. Did you feel **joint pain** in the past 3 days? | 0 = No<br>1 = Yes |
| 9.8.1. **If yes:** How bad was the joint pain? | 1 = The joint pain did not interfere with my activities<br>2 = The joint pain somewhat interfered with my activities<br>3 = The joint pain was considerable and prevented my daily activities |
| 9.9. Did you have **muscle aches** in the past 3 days? | 0 = No<br>1 = Yes |
| 9.9.1. **If yes:** How bad were the muscle aches? | 1 = The muscle aches did not interfere with my activities<br>2 = The muscle aches somewhat interfered with my activities<br>3 = The muscle aches were considerable and prevented my daily activities |
| 9.10. Did you feel **tired (fatigued)** in the past 3 days? | 0 = No<br>1 = Yes |
| 9.10.1. **If yes:** How bad was the tiredness? | 1 = The tiredness did not interfere with my activities<br>2 = The tiredness somewhat interfered with my activities<br>3 = The tiredness was considerable and prevented my daily activities |

***Cohort post-vaccine follow-up questionnaire (all participants)***

| **Questions to be automatically populated/ completed by study team** | **Response value/ coding or type** |
|---|---|
| 10. Participant unique identifier (from baseline questionnaire) | Unique Numeric/ text/ alphanumeric |
| 11. Identifier of the follow-up questionnaire | Unique Numeric/ text/ alphanumeric |
| 12. Questionnaire completed? | 0 = No<br>1 = Yes |
| 12.1. **If no:** Reason why follow-up questionnaire was not completed | 0 = No reason given<br>1 = The study takes too much time<br>2 = Not interested anymore<br>4 = Death<br>5 = Participant unreachable<br>6= Other (Specify) |
| 12.1.1. **If death**: Reason for death | Text |

*Questions that are shaded grey will not be included in the USSD-based questionnaire. Study staff will call participants answering ‘yes’ to hospitalization or COVID-19 diagnosis to collect the additional information in the grey-shaded sections.

| **Questions to be completed by interviewing the participant via USSD-based or phone call-based questionnaire*** | **Response value/ coding or type** |
|---|---|
| 13. How many times did you seek medical care between date X and date Y? (e.g.at a local health centre or hospital) | 0 = Did not seek medical care<br>1 = 1 times<br>2 = 2 times<br>3 = 3 times<br>4 = Other (Specify) |
| 14. Were you hospitalized since **[**date of last contact**]?** | 0 = No<br>1 = Yes |
| 14.1. **If yes**: What was the date of hospital admission? | dd/mm/yyyy |
| 14.2. Have you been discharged from hospital? | 0 = No<br>1 = Yes |
| 14.3. What was the date of hospital discharge? | dd/mm/yyyy |
| 14.4. What was the reason for your hospitalization? | Text |
| 14.5. What was the diagnosis by the healthcare provider? | Text |
| 14.6. Picture/ attachment of discharge report available? | 0 = No<br>1 = Yes |
| 14.7. Name and place of hospital | Text |
| 15. Were you diagnosed with COVID-19 by a healthcare professional? | 0 = No<br>1 = Yes |
| 15.1. **If yes**: Was the diagnosis based on a laboratory test? | 0 = No<br>1 = Yes<br>2 = I do not know |

| | |
|---|---|
| 15.2. What was the date of symptom onset? | dd/mm/yyyy |
| 15.3. Was admission to the intensive care unit necessary? | 0 = No<br>1 = Yes |
| 16. Are you pregnant? | 0 = No<br>1 = Yes<br>2 = Not applicable |
| 17. Have you received another COVID-19 vaccine dose since last contact? | 0 = No<br>1 = Yes |
| 8.1. **If yes**: Dose of **current** COVID-19 vaccine? | 1 = 2nd vaccination<br>2 = 3rd vaccination<br>3 = 4th vaccination<br>4= other (specify |
| 8.2. Date of **current** vaccination | dd/mm/yyyy |
| 8.3. Vaccine brand/manufacturer | 1= AstraZeneca<br>2= Pfizer<br>3= Johnson & Johnson<br>4= Moderna<br>5= Sputnik V<br>6= Sinopharm<br>7= Other (specify<br>8= Don't know |

***Reactogenicity: post-vaccine reactogenicity questionnaire (reactogenicity subset only)***

*Questions that are shaded grey will not be included in the USSD-based questionnaire. Study staff will call participants answering 'yes' to any of the reactogenicity events to collect the additional information in the grey-shaded sections.

| **Question*** | **Response value/ coding or type** |
|---|---|
| 18. Participant unique ID (from baseline questionnaire) | Unique Numeric/ text/ alphanumeric |
| 19. ID for the record of post-vaccination reactogenicity corresponding to each date | Unique Numeric/ text/ alphanumeric |
| 20. Date of record | dd/mm/yyyy |
| 21. Did you feel **feverish** yesterday? | 0 = No<br>1 = Yes |
| 21.1. **If yes:** Did you measure your temperature yesterday? | 0 = No<br>1 = Yes |
| 21.2. **If yes:** What was your temperature? | 1 = Below 38.0°C (below 100.4°F)<br>2 = 38.0°C to 38.4°C (100.4°F to 101.12 °F)<br>3 = 38.5°C to 38.9°C (101.3°F to 102.02°F)<br>4 = Higher than 39.0°C (higher than 102.2°F)<br>5= I can't remember |
| 21.3. How did you measure the temperature? | 1 = Oral (in the mouth)<br>2 = Rectum (in the anus)<br>3 = Armpit<br>4 = Ear<br>5 = Forehead |
| 22. Did you feel **like vomiting (nausea)** yesterday, or did you vomit? | 0 = No<br>1 = Yes |
| 22.1. **If yes:** How severe was the nausea/ vomiting? | 1 = The nausea/vomiting did not interfere with my activities<br>2 = The nausea/vomiting somewhat interfered with my activities<br>3 = The nausea/vomiting was considerable and prevented my daily activities |
| **23.** Did you experience **a feeling of weakness or not feeling well (malaise)** in the past 3 days? | 0 = No<br>1 = Yes |
| 23.1. **If yes:** How severe was the malaise? | 1 = The malaise did not interfere with my activities<br>2 = The malaise somewhat interfered with my activities<br>3 = The malaise was considerable and prevented my daily activities |

| Question* | Response value/ coding or type |
|---|---|
| **24.** Did you have **a feeling of being cold (chills)** yesterday? | 0 = No<br>1 = Yes |
| 24.1. **If yes:** How severe were the chills? | 1 = The chills did not interfere with my activities<br>2 = The chills somewhat interfered with my activities<br>3 = The chills were considerable and prevented my daily activities |
| **25.** Did you have a **headache** yesterday? | 0 = No<br>1 = Yes |
| 25.1. **If yes:** How bad was the headache? | 1 = The headache did not interfere with my activities<br>2 = The headache somewhat interfered with my activities<br>3 = The headache was considerable and prevented my daily activities |
| **26.** Did you feel **joint pain** yesterday? | 0 = No<br>1 = Yes |
| 26.1. **If yes:** How bad was the joint pain? | 1 = The joint pain did not interfere with my activities<br>2 = The joint pain somewhat interfered with my activities<br>3 = The joint pain was considerable and prevented my daily activities |
| 27. Did you have **muscle aches** yesterday? | 0 = No<br>1 = Yes |
| 27.1. **If yes:** How bad were the muscle aches? | 1 = The muscle aches did not interfere with my activities<br>2 = The muscle aches somewhat interfered with my activities<br>3 = The muscle aches were considerable and prevented my daily activities |
| 28. Did you feel **tired (fatigued)** yesterday? | 0 = No<br>1 = Yes |
| 28.1. **If yes:** How bad was the tiredness? | 1 = The tiredness did not interfere with my activities<br>2 = The tiredness somewhat interfered with my activities<br>3 = The tiredness was considerable and prevented my daily activities |
| 29. Questionnaire completed? | 0 = No<br>1 = Yes |

| Question* | Response value/ coding or type |
|---|---|
| **12.1 If no:** Reason why follow-up questionnaire was not completed | 0 = No reason given<br>1 = The study takes too much time<br>2 = Not interested anymore<br>4 = Death<br>5 = Participant unreachable<br>6= Other (Specify) |
| 29.1.1. **If death:** Reason for death | Text |
